# YeZzzs Does it: Studying Sleep and Emotion using the Digital Rest-Activity Rhythms of Kanye West’s Tweets

**DOI:** 10.1101/2025.02.03.25321602

**Authors:** Matthew J. Reid, Darlynn M. Rojo-Wissar, Michelle Mei, Moira Differding, Michael T Smith, Michael G Smith

## Abstract

One of the greatest challenges faced by the field of precision medicine is the identification of biomarkers capable of detecting clinically meaningful change at the individual level, not just amongst large-scale population studies. To this end, the evermore present social mediaverse provides unparalleled access to ecologically-valid databases of digital-biomarkers, that could be leveraged with single-user precision to support mental health care by monitoring use-patterns and emotional state. Despite this potential, investigation into how social-media use can be used to study sleep-wake behaviors has been remarkably scant, in part due to a lack of established methods to detect and estimate sleep using social-media activity. We present here a new approach to using social media-based data to track both sleep and mood, with potential applications to mental health monitoring and prevention. Amongst demonstrating proof of concept, we also provide an ethical and theoretical framework of how to proceed amongst this sensitive but potentially highly fruitful field.

## Background

One of the greatest challenges faced by the field o f p recision m edicine i s t he identification o f b iomarkers c apable o f d etecting clinically meaningful change at the individual level, not just amongst large-scale population studies. To this end, the evermore present social mediaverse provides unparalleled access to ecologically-valid databases of digital-biomarkers, that could be leveraged with single-user precision to support mental health care by monitoring use-patterns and emotional state. An illustrative study[1] published in Science demonstrated the presence of circadian rhythmicity at the large (n=2.4mn) population level in the emotional content of ‘Tweets’ with positive emotionality declining, and negative emotion increasing with progression of the day. These findings a lign w ith e xperimental w ork in humans measuring biological markers of circadian rhythm, suggesting these naturalistic assessments have potential to represent credible biomarkers. Despite these early advancements, further investigation into how social-media use can be used to study sleep-wake behaviors has been remarkably scant, in part due to a lack of established methods to detect and estimate sleep using social-media activity.

Case studies of high-volume Twitter users represent a means through which to validate novel techniques in homogenous longitudinal data. In an elegant effort, a previous study by Ronnenberg[2] used Tweets to estimate sleep-opportunity by analysing the circadian rhythmicity of a single social media account ( 6.5hrs in the case of Donald Trump). However, links between these digital rest activity rhythms (dRARs) and clinically or socially relevant indices, such as the emotional content of tweets, were not examined, potentially due to the complexities involved in integrating these dRARs with the highly complex yet rich nature of tweet contents. Extending this work, we developed a methodological process to examine the relationship between a user’s sleep-wake pattern and the daytime emotional content of their tweets. As these tools are ultimately intended for use at the level of the individual user, here we present proof-of-concept data from a single user to identify periods of sleep disturbance, based on a user’s habitual sleep window and evaluated their impact on next-day emotional state. In the present analysis we examined the rhythmicity and content of tweets from a single high-volume user to identify when activity occurred during a user’s habitual sleep window and evaluated impact on daytime emotional state. can be helpful to the reader.

## RESULTS

Our model confirmed findings[1] at the population level that tweets fluctuate significantly across the 24-hour cycle with positive emotion declining with the progression of the day (P <0.001), whilst negative emotion increased and peaked towards the evening (P <0.001) (See Figure 1). Next, we tested the effect of nocturnal tweeting on next-day emotional tweet content whilst controlling for time-of-day of the daytime tweets, month, and the tweet-word count, to mitigate their potential confounding. Our models demonstrated that increased nocturnal tweeting was significantly associated with negative emotional content of tweets the following day (Bonferroni-corrected P = <0.001) and was also linked to significantly reduced positive emotion the following day (Bonferroni-corrected P = <0.001). When examining tweets across the previous week, increased nocturnal tweeting was also significantly associated with decreased positive emotional content (Bonferroni-corrected P = 0.041), but not with negative emotional content (Bonferroni-corrected P= 0.432)

**Figure 1.**
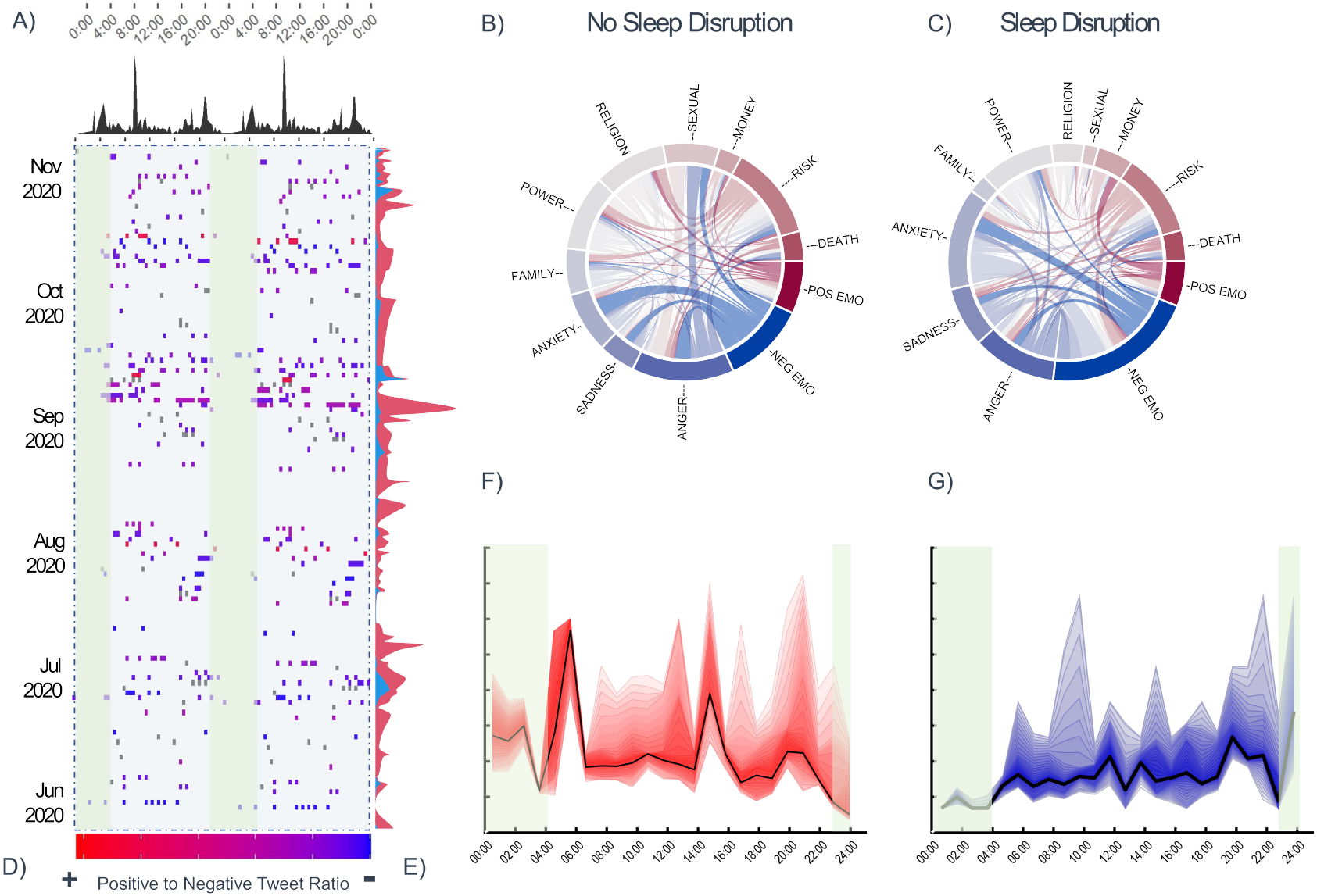
Visualisation of emotional tweet content: A) Daily tweet frequency in 20-minute bins across the 24h day, averaged from 258 days of Twitter data and 1,868 tweets. B) and C) The semantic network of principally weighted emotional constructs during days which were preceded by known nocturnal tweeting, versus those in which no nocturnal tweeting was present. D) Double plot: visualises the relative emotional ratio of positive to negative content of tweets in each 20-minute bin across the 24 hour day across a representative sample period from June to November 2020, with red indicating more positive content, blue more negative content, and purple about the same amount of positive and negative content (see color bar key on horizontal axis). Grey squares indicate the presence of tweets which were timestamped but were omitted from sleep-linguistic analyses. E) Presented along the right-vertical axis of D, further depicts the frequency of positive (in red) tweet content overlaid with the frequency of negative (in blue) tweet content per day across the assessment period. F) and G) positive and negative valences of tweets when plotted as a function of the average 24h period. As positive emotion was much more prominent than negative emotion, the y axis depicts relative units to facilitate comparisons of circadian trajectories, as opposed to relative magnitudes. Shaded error bars indicate 95 percent Confidence Intervals of the relative values. Note. The nocturnal period (L5 window) from 23:00-04:00 is shaded in light green.)

These data show that previous nights’ nocturnal tweeting is associated with a decrease in positive Tweet content the following day (defined as the period outside the L5 window: 23:00-04:00), whilst negative tweet content remains stable. These observations align with previous social media studies1, and causal sleep-disruption studies4,6 in humans which report substantial blunting of positive emotion and modest or no increases in negative emotion, suggesting our methods may be sensitive to detecting both sleep-determined emotional-state and circadian effects in emotion. Broadly, these findings give credence to the notion that sleep disturbance results in a generalised ‘negative emotional-bias’ even when assessed through naturalistic observations of a single user. Nevertheless, it should be noted that whilst dRARs do allow us to estimate maximal sleep opportunities, sleep cannot be definitively inferred from nocturnal digital quiescence.

## DISCUSSION

### Potential applications of dRARs in mental health care

Our methods have the advantage of leveraging real-world data which reflects an individual’s un-prompted and spontaneous thoughts and feelings, and are sensitive to change on this individual level, making them ideal candidates for deployment as digital biomarkers, and mHealth interventions. Although continuous monitoring of social media content poses numerous ethnical concerns, when applied benevolently, automated text-mining algorithms could have the potential to be used to identify individuals susceptible to emotional distress due to circadian and sleep-related disruption, which could facilitate the administration of ‘Just-In-Time Adaptive Interventions7’, or signpost to appropriate support. These may complete or even integrate with automated harm-prevention mechanisms present on social media sites, such as ‘pop-ups’ offering support resources whenever a user’s activity is flagged due to the use of keywords associated with suicide.

### Methodological Challenges

Much in the same manner that wearable devices which track sleep must be compared against gold-standard metrics of sleep, examinations of how sleep estimates from dRARs align with predetermined benchmarks for accuracy must be conducted, by assessing their synchronicity with well-defined biological (e.g., circadian), environmental (e.g., night/day or light/dark) or social rest-activity (e.g., work-leisure) rhythms when measured using techniques commonly held as accurate assays of these rhythms. For instance, determination of the peak of dRARs activity relative to one’s dim light melatonin onset (DLMOn) peak could provide a window into alignment with, or equally orthogonality, endogenous biological cycles. Similarly, comparisons with 24-hour actigraphy, would represent a logical follow-up in this validation pipeline. From a translational perspective, future studies may consider assessment of clinical validity amongst known ‘at-risk’ populations as all the lexicons used in NLP methods such as sentiment analysis and LIWC are based on healthy individuals. Once the goal becomes the identification of language indicative of clinically-significant emotional distress, to what extent can we consider these normative datasets to be valid. We recommend the development of lexicons based on distinct clinical populations (e.g., MDD, GAD, PTSD, BPD), which may maximize the sensitivity of these models. Moreover, language by nature differs drastically amongst cultures, and societies. Therefore, it is important to consider whether models trained using one particular language be transferred to another, as well as whether these models perform equally well across different populations. Equally important is the question of how differences in language are perceived across different ethnic identities, races, and cultures. Given that certain cultures, ethnicities and genders communicate emotion much more readily than others, therefore how conspicuous should language be before it is considered indicative of emotional distress. Much in the same way that disorder-specific models may contribute towards increased precision for the detection of emotional distress, models which adapt to the differences in sleep behaviours (e.g., bi-phasic sleeping or siesta cultures) may prove beneficial.

Perhaps the most conspicuous methodological challenge associated with deriving rest-activity rhythms from social media use is the continuity and completeness of the data. Thorough sensitivity analyses to determine both the minimum amount of data required to accurately determine rhythms, we as well as the precision point at which further data brings no further gain in model accuracy should be determined, to ensure the credibility of predictions, as well as protection against unnecessary exposure of data when it is not necessary to inform models. Although the present study focused exclusively on social media activity as its data-source, it should be noted that the proposed methods are by no means limited to these data. For better or worse, activities of daily living increasingly rely on the use of smartphones or mobile applications8,9, all of which have potential to inform models used to train predictive dRARs. Moreover, a significant portion of individuals report using their phone immediately before going to bed (e.g., checking social media or emails, setting alarms, plugging a phone into a charging socket) and immediately again upon awaking (e.g., to cancel an alarm).10,11 Triangulating data from multiple digital sources within a user’s digital device network (e.g., app usage, screen inactivity) could therefore, have the potential to yield a remarkably accurate digital fingerprint of the user’s bed and rise time patterns, over time helping to build a highly intelligent model of the user’s dRARs, and flag any deviations from normative cycles.

### Tending to the ethical and legal landscape of social media monitoring

Although a great deal of research and unknowns stands between the present and any form of real-world implementation, earnest exploration of the ethical and legal landscape of such advancements is certainly warranted. Principally, consideration of how the use of social media data to monitor an individual’s sleep behaviors of emotional well-being aligns with the basic ethical principles upheld by medicine requires addressing. For instance, where does the responsibility of technology devised to monitor health behaviours and emotional wellbeing end? In the process of monitoring emotional word content, do we also uphold a responsibility to monitor other behaviours which may be perceived as threatening (e.g., abuse, terrorism) to others. One could easily draw comparisons with analogues amongst the medical field, such as incidental findings of structural abnormalities which occur during MRI scans. Assuming consent is deemed a requirement, the stage and source at which consent is obtained presents another challenge. For instance, should this take place at the terms and conditions level of the end-user platform, in essence permitting monitoring of all users, or should there be an independent opt-in process external to these initial terms and conditions, either allowing users to opt-in or opt-out. Once entertaining the realms of informed consent, the age at which one is considered competent of consent or assent becomes an immediately pertinent manner. Most social media platforms permit adolescent users, typically below the age of what most healthcare systems would be considered medically autonomous. Should these users be omitted from monitoring on the grounds of inability to consent, or should consent be sought through legal guardians? Whilst ethically straightforward and defensible on the grounds of consent, omitting adolescents and those under the age of legal consent entirely from the process has potential to create an age-group disparity in the availability of preventative health measures. One must also of course take into account the fact that younger users are those most likely to be high-volume users, those most likely to convey emotional distress on social media, as well as those at greatest risk for sleep and circadian disruption as well as the development of preventable mental illness. Therefore, perhaps, the benefits may out-weight the risks in this age group.

## CONCLUSION

We demonstrated here the feasibility of methods which could be used to leverage the digital rhythms of social media user’s (dRARs) for the benefit of monitoring sleep and emotional wellbeing. Future work requires a multidisciplinary approach through parallel lines of investigation spanning scientific, digital, medical, and philosophical disciplines. The work we present here represents a modest step in the roadmap we outline in this paper, and we outline several recommendations for key questions to facilitate and stimulate future work in this arena. Importantly, several vital questions with more provocative and wide-reaching implications need to be overcome in order to bridge the gap between proof-of-concept and implementation.

## Data Availability

data is derived from publically available social media data

## AUTHOR CONTRIBUTIONS

M.J Reid had the original idea for the study and performed the analyses. D.M Rojo-Wissar tried to talk him out of it. M. Differding reluctantly helped with the analyses, and in a similar manner M Mei. somewhat hesitantly assisted with the data-visualisation. Both M.Smith’s conveniently shared the same name, but also provided editorial assistance and senior guidance.

**Dr Matthew J. Reid BMedSci MSc Dphil** Department of Psychiatry and Behavioral Sciences, Johns Hopkins School of Medicine. Contact him/her at Department of Psychiatry and Behavioral Sciences, Johns Hopkins School of Medicine. Contact him/her at

## METHODS

1,868 ‘Tweets’ were extracted from the user-profile (available at www.twitter.com/@Ye“) across 258 days using the Twitter.com Application Programming Interface (API) and a custom programming script written in Python. To approximate nocturnal from diurnal tweets, data were mean-averaged across the period, and a sliding window function applied to determine the least active 5 hours (L5 index) of the 24-hour period, a common method used to determine the nadir of circadian quiescence3 and how we defined the “nocturnal” window. We subsequently identified days in which tweets were preceded by Twitter activity during the previous nocturnal window and derived the number of tweets during that window, as well as the total number of nocturnal tweets over the previous seven days (see supplement for detailed methods). In accordance with the previous study1 which explored circadian aspects of emotional tweet content, we also derived measures of positive and negative affect for each tweet using a natural language processing model (Linguistic Enquiry and Word Count [LIWC5] analysis), using the ‘posemo’ and ‘negemo’ output variables which give an overall positivity and negativity score for each tweet based on a validated linguistic dictionary of 6,400 words. We subsequently calculated mean positive and negative affect across hourly and monthly bins to assess circadian and seasonal effects. Due to the large number of possible variables produced by LIWC, we opted to limit our model to the indices of positive and negative affect. Models were conducted with zero-inflated Poisson regression controlling for time and month of each tweet to prevent confounding. In separate sensitivity analyses, we also tested the relationship of time and month with positive and negative emotion using zero-inflated Poisson regression (testing for main and interaction effects of hour, month, and affect (positive vs negative) both to serve as a validation check for their pre-specified inclusion as covariates, and to test the sensitivity of the model to detect known effects observed at the population level in a single user. Finally, we visualized data using a double plot, adapted from a common method used in circadian actigraphy research, along with semantic networks for the emotional constructs identified by the LIWC analysis (figure 1). Tweets were extracted from the twitter page between the dates (04/14/2018) – (11/04/2020) as these dates spanned the range of the earliest and latest tweets available. Prior to linguistic analysis text data were cleaned to remove special characters which may influence algorithmic outputs. ‘re-tweets’ and tweets without any linguistic content (such as images/’gifs’/’memes’) were included as time-indicative data-points, but were omitted from linguistic analysis, due to the presence of linguistic content from other users. In other words, time data from re-tweets images etc. were used to inform when tweeting activity occurred, but were not included as observations of outcome variables in the statistical models. Character symbols conveying emoticons were re-coded into lexical equivalents, according to of industry-standard dictionary (Emoji List, v15.0 (unicode.org)) containing the accepted nomenclature assigned to respected UTF-8 encoded characters (e.g. ‘:)’ = ‘slightly smiling face’). Temporal Data Analysis: Timestamps were first converted from native format (UTC) to local time (e.g. PDT, PST, EST) before any temporal analyses. To estimate the period of peak quiescence purported to represent the user’s sleep window we adopted a common and well-validated approach used in assessments of human circadian rhythms, by calculating the the least active 5 hours of each day (the L5 index). To achieve this, we pooled the data and averaged it into one 24hour period. A sliding window function with a step size of 1 hour was then used to calculate the total number of tweets which occurred within that 5-hour window. We permitted ‘wrapping’ of this window to prevent an assumed to be stationary 24-hour period from identifying false L5 estimates. For example, the following L5 window: 20:00, 21:00, 22:00, 23:00. 0:00 is stationary as each of the 1hour bins falls within the bounds of one ‘finite’ 24-hour cycle. However, the L5 window: 23:00 —00:00 — 01:00 — 02:00 — 03:00 falls across the boundaries of two. An ‘unwrapped’ sliding window would identify this single period as two independent activity troughs (23:00-00:00 , 00:00-04:00) biasing the estimated L5. Instead, we permitted ‘wrapped’ L5 windows which spanned between two 24-hour periods. We calculated L5 in our data as the 5-hour window (in this case 23:00-04:00) which contained the lowest total number of tweets, when averaging across all days. Next, we assigned a binary classifier to each tweet according to whether the tweet was sent within or without the L5 window. For the purposes of interpretation and reporting, we defined tweets within the L5 window as ‘nocturnal’ and ‘diurnal’, and it should be noted that these labels do not necessarily indicate the position on the dark/light cycle. Nevertheless, the primary geographical latitude of the user (mainland USA, primarily west coast) resulted in this window always occurring during hours of darkness. These binary labels (0, 1) were then summed to create the total number of nocturnal/L5 tweets. Although many users log daily activity on social media platforms, use is sporadic and non-contiguous in nature, and daily use can neither be assumed nor expected from a given user. Therefore, we also calculated a metric representing the total number of tweets over the preceding seven days which occurred during the L5 period. This allowed us to maximize statistical power by increasing data points, and also to test the clinically relevant yet hypothetical influence of accumulated hypothetical sleep/circadian disruption on mood. Linguistic Analyses The most frequently adopted approach to study Twitter and other social media content is sentiment analysis. Sentiment analysis offers a low dimensional representation of emotional language by integrating several complex linguistic concepts and word valences into one measure of relative positive/negative emotion, ranging from –1 (negative) to 0 (neutral), to +1 (positive). However, whilst this may be appropriate for decoding emotionality in most instances, such an approach conflates positive and negative emotion into a single dimension, assuming their inter-dependence. This assumption departs from the substantial body of literature in the sleep field which indicates that positive and negative emotion are independent/orthogonal dimensions that are differentially affected by sleep loss and circadian trajectories. Therefore, in accordance with a robust seminal study1 which explored circadian aspects of emotional tweet content, we also derived measures of positive and negative affect for each tweet using Linguistic Enquiry and Word Count (LIWC2) analysis, by using the ‘posemo’ and ‘negemo’ output variables which give an overall positivity and negativity score for each tweet based on a validated linguistic dictionary of 6,400 words. We subsequently calculated mean positive and negative affect by averaging across hourly and monthly bins to assess circadian and seasonal effects. Due to the large number of possible variables produced by LIWC, to reduce alpha inflation we opted to limit our analyses a priori to the outcomes of positive and negative affect. Primary analyses tested for a relationship between the number of nocturnal tweets and emotion (positive, negative). Data visualization In order to facilitate interpretation of the data generated by our methods to the most commonly used benchmark technique in sleep and circadian science, we produced circadian ‘double plots’ to visualize the bins of activity over the entire period in 48-hour epochs (rows), displayed across time (columns). Whilst Ronenberg used a dedicated circadian software, we opted for an approach that would facilitate more open-source development of these methods. In order to do so, we adopted an approach akin to those used to visualise other time-series sleep/circadian data generated by actigraphy and EEG. Such approaches use continuous time series of either binary (activity/no activity) or continuous (activity count/epoch) assigned to each predefined time bin/epoch. We opted for a 20minute epoch, as the results of our initial sensitivity analyses resulted in this epoch lengths of 20minutes generating the highest visual and temporal resolution, whilst still permitting clear visual delineation of temporal fluctuation Rythmns. 20 minutes are also frequently used as meaningful epochs in studies employing activity methods which facilitates meaningful comparison. Whilst a standardized consensus epoch-length may ultimately be reached following further investigations, we recommend an iterative approach towards tailor epoch length to each specific population/participant based on the nuances of the observed data, and factors such as tweet resolution (e.g. the number per hour), the length of the total time period and the overall tweet frequency and total numbers of tweets. The localtime timestamps of the Tweets were first down sampled using the MATLAB Signal Processing Toolbox to assign replace the timestamp with the time-epoch in which it fell. Epochs repeated as follows between 00:00 and 11:59 for each day in which tweets were present: hh:00-hh:19 — hh:20-hh:39 — hh:40-hh:59. In order to then epoch the tweet times, we then simulated a time-series of 20-minute epochs across the entire period (total 705,600 bins). The time series was transposed into vertical format, and the rows of tweet data were joined using a left join function so that the timestamps of the tweet were matched to their appropriate bin, and intermediate periods were left empty. We then visualised the data as a 48hr double plot using R, by using a heatmap function, where each cell in the heatmap matrix represented a 20min time bin, and was filled if Tweet activity occurred during that bin. A color bar was used to represent a continuous value representing the ratio between positive and negative tweets. Dark blue indicated a tweet ratio of 1:0 negative to positive, where as bright red indicated 0:1 negative to positive. Although our analyses conceptualized these two dimensions as orthogonal concepts, we opted for this approach during data visualization as a compromise to using two different double plots to present negative and positive emotion independently, whilst also indicating overall tweet (non-emotional) activity. Data which were timestamped, but not used in analyses (e.g. retweets) were indicated by grey shading in the matrix. In addition to this double plot, we also visualized trajectories of positive and negative tweets by plotting the posemo and negemo values averaged across the entire period. Finally, although our a priori analyses were limited to the posemo and negemo values generated by the LIWC analysis, the outputs of the LIWC algorithm provide a much more granular picture of emotionality across the spectrum of both positive and negative emotions. In order to represent these relationships visually, without generating further hypotheses or statistical tests, we conducted network analyses to visualise the relationship among nodes, We selected the 10 emotional constructs with the highest weightings assigned by the LIWC algorithm (Anger, Sadness, Anxiety, Family, Power, Religion, Sexual, Money, Risk, Death) and established a network matrix combining these values with positive and negative emotion. The relationship (signified by the thickness of the cord) between nodes was plotted using cord plots, and the relative weighting of the emotion categories was indicated by the relative proportion of the circumference band.

Statistical Analyses and Diagnostics We tested the hypothesized relationship between nocturnal tweeting and daytime emotion in relation to the previous-nights nocturnal tweeting and average number of nocturnal tweets for the seven previous days. Analyses of positive and negative emotional words specified the inclusion of time of day, month, and the total number of words in the tweet a priori, to prevent confounding their confounding. Nevertheless, we first tested the relationship of these confounds with positive and negative emotion using zero inflated generalised linear modelling (testing for main and interaction effects of hour, month, and affect (positive vs negative) both to serve as a validation check for their pre-specified inclusion in models, and to test the sensitivity of the model to detect known effects observed at the population level in a single user. Tweets for which there was no data available the previous day, or previous seven days were excluded from the respective analyses. We hypothesized that increased tweeting during the nocturnal period would be associated with decreased positive and increased negative daytime affect (see detailed hypotheses below). We first probed the central tendency and distributions of our independent and dependent variables using histograms and statistical diagnostic tests. The Shapiro-Wilk test indicated that the proposed predictors (Number of nocturnal tweets: last night and past week) and outcomes deviated significantly from a normal distribution (both p ¡0.001). Similarly, the Anderson-Darling test confirmed the departure from normality for each variable (both p ¡0.001). Further-more, positive emotion, negative emotion, frequency of nocturnal tweets (both daily and weekly), exhibited substantial skewness (¿+2 or ¡-2) and kurtosis (¿+3 or ¡-2) values, suggesting outcome and predictor variables followed a log-normal distribution with right-skewing in the presence of zero values. As a result, we employed a zero-inflated Poisson model to account for the excess zeros in the data and appropriately handle both normally distributed predictors and non-normally distributed outcomes. These methods best handle the logarithmic distribution and are robust to deviations from the assumption requirements of linear models., The zero-inflated Poisson model was implemented using the Rigby and Stasinopoulos method facilitated by the use of the the GAMLSS (Generalized Additive Models for Location, Scale, and Shape) framework in the R programming language. We controlled the fitting process using 500 cycles to improve the convergence and stability of the model estimation. We evaluated model performance by examining the global deviance, reflecting overall goodness-of-fit, as well as the Akaike Information Criterion (AIC), and the Schwarz Bayesian Criterion (SBC) values, reflecting model parsimony, with lower values indicative of better trade-off between model complexity and fit. Finally, we examined overdispersion, which we calculated as the ratio of the residual deviance to the residual degrees of freedom. Based on our prior work, we hypothesised that positive and negative emotion values would demonstrate relatively weak levels of correlation, and therefore proposed independent univariate models a priori. However, during the data-cleaning phase we verified (pre-analysis) whether univariate or multivariate modelling may be most appropriate by calculating the correlation between outcome variables, which demonstrated weak correlation (r = -0.052, p ¿ 0.05) and therefore weak interdependence. This confirmed our assumption, and justified the use of univariate models which permit the exploration of unique associations between each outcome variable and the predictors without assuming dependency between the two.

Our data relied on samples obtained from a single participant. Therefore, in order to assess the stability and variability of the coefficients estimated by the GAMLSS model, a bootstrapping procedure was performed. Bootstrap is a resampling method which involves randomly drawing samples with replacement from the original dataset. For each bootstrap sample, a GAMLSS model was fitted with the same set of predictors and response variable. The bootstrapping process was repeated multiple times, resulting in a distribution of coefficient estimates for each predictor which provide a measure of the uncertainty associated with the coefficient estimates. The results showed that the coefficients for the predictors month simple, WC, LocalTimehhEX, and total noct tweets week were reasonably stable, with narrow confidence intervals. To ensure we obtained the most accurate estimations of our model’s validity our model stability estimates, we opted for a much higher than usual number of bootstrapping iterations (20,000). However, a standard computing system may lack the computing power to run such models, therefore future investigators may wish to consider the balance between model stability and computational power.

